# Neighborhood socioeconomic status and environmental burden show diverging associations with cognitive impairment in community-dwelling older adults

**DOI:** 10.64898/2026.10.13.26365488

**Authors:** Kevin Shen, Kyra M. Bonta, Meseret K. Alemu, Emma Corbett, Beniam D. Darge, Lauren K. Hilderbrand, Kaitlyn Richardson, Isabella M. Blum, Stephanie S. Ogonuwe, Madison K. McGuire, Diya H. Pandhi, Lewis W Gallagher, Jennifer M. Hunter, Fredrica Grant, Jackiethia Butsch, Neale Davis, Wy’Esha Simmons, Loletia Davis, Pamela Shields, Sue Cooper, Missy Rouse, Kimberly S. Wilson-Lawson, Carlotta Hughes-Culp, Mary Howard, Mertice Monroe, Matthew Coleman, De’leah Leath, Jimmy Henry, Pamela Gregory, Douglas W. Scharre, Jeremy G. Grant

## Abstract

**OBJECTIVE:** Research on neighborhood context and cognitive health often relies on composite indices that collapse neighborhood socioeconomic status (SES) and aspects of the built environment into a single score. This study examined the extent to which neighborhood SES and built environment were associated with late-life cognitive impairment.

**METHOD:** Participants were 281 predominantly Black older adults who completed the Self-Administered Gerocognitive Exam (SAGE), a brief cognitive screening measure. Participants also provided their home address to obtain four neighborhood metrics: Two composite measures of neighborhood disadvantage, the Environmental Justice Index (EJI) and the EJI Social-Environmental Rank, as well as the EJI Social Vulnerability Rank as a measure of neighborhood SES, and the EJI Environmental Burden Rank to measure exposure to hazardous aspects of the built environment.

**RESULTS:** Lower neighborhood SES was associated with higher odds of cognitive impairment. In contrast, greater environmental burden was associated with lower odds of impairment, with years of education as a moderator; the association was strongest among individuals with high levels of education. The two composite scores were not associated with cognitive impairment.

**CONCLUSION:** Composite measures of neighborhood features may obscure distinct pathways of their effects on cognition.

## INTRODUCTION

Early detection of cognitive decline is a cornerstone of effective care for individuals with Alzheimer’s disease and Alzheimer’s disease-related dementias (AD/ADRD). Identifying impairment in its earliest stages is essential for distinguishing underlying etiologies, optimizing treatment options, and offering opportunities to address modifiable dementia risk factors. A wealth of research has demonstrated that a substantial portion of dementia cases worldwide could be reduced by addressing modifiable risk factors, such as cardiometabolic disease, depression, hearing loss, vision loss, and other chronic health conditions across the lifespan (Livingston et al., 2024) Importantly, the determinants of late-life cognitive health extend well beyond the individual. Although research on cognitive aging has historically emphasized individual-level factors such as education, medical conditions, and health behaviors, neighborhood environments form a pivotal but comparatively understudied social determinant of brain health (Finlay et al., 2022). In comparison to older adults living in affluent neighborhoods, living in a disadvantaged neighborhood has been linked to poorer executive function (Hunt et al., 2021), faster cognitive decline and progression to dementia (Vassilaki et al., 2022), and lower cerebral volume and hippocampal volume, even among cognitively unimpaired older adults (Hunt et al., 2020; Powell et al., 2020).

As defined by the CDC’s Healthy People 2030 framework, there are at least two dimensions to consider when examining the influence of neighborhood conditions on health outcomes: (1) neighborhood socioeconomic status (SES) and (2) the built environment (Office of Disease Prevention and Health Promotion, n.d.). Neighborhood SES examines the extent to which a particular residential area is economically advantaged or disadvantaged, including the level of educational attainment, employment, home ownership, vehicle ownership, and housing quality averaged across its residents. Poorer neighborhood SES has been linked to poorer cognition (Vassilaki et al., 2022; C. G. Wong et al., 2023) and greater risk of cognitive decline and dementia (Coogan et al., 2025; Dintica et al., 2023; Li et al., 2026). However, dementia is also often underdiagnosed among people living in areas of low neighborhood-level SES (Godard-Sebillotte et al., 2025; Holm et al., 2022). On the other hand, the built environment refers to aspects of a neighborhood that have been modified by human activities to provide infrastructure for daily life. Some features of the built environment foster healthy cognitive aging by enabling physical movement and expanding access to health-promoting activities (Chan et al., 2023), such as living in a neighborhood with adequate greenspace (L. Besser, 2021; Pescador Jimenez et al., 2026) or gathering spaces that foster physical activity (Finlay et al., 2021) and social connection (Finlay et al., 2020). Conversely, comparatively fewer studies have investigated how *environmental burden*—exposure to hazardous aspects of the built environment—may harm late-life cognitive health. The most well-studied aspect of environmental burden is air pollution, which has been linked to increased dementia risk (Shi et al., 2023; Weuve et al., 2021) and appears to mediate the risks associated with other aspects of the built environment. For example, greater exposure to heavy-traffic transportation infrastructure has been associated with increased incidence of Alzheimer’s disease and other neurodegenerative disorders, likely due to air pollution (Chen et al., 2017). Furthermore, exposure to certain heavy metals has been associated with increased dementia risk (Killin et al., 2016), which may affect individuals living near toxic waste sites.

Most research on the relationship between neighborhood context and dementia risk relies on composite place-based indices that aggregate socioeconomic and environmental features into a single summary score the implicit assumption is that their components align. This presents a measurement challenge that has not been widely studied in cognitive aging research. Neighborhood SES and the built environment do not necessarily act on cognitive health and dementia risk in the same direction; living in a high SES neighborhood does not guarantee a favorable physical environment or a lack of exposure to environmental pollutants, and the reverse can also hold true (Wagner et al., 2025). If socioeconomic and environmental components relate to cognition in divergent directions, a composite rank that sums them may obscure both associations and yield a null or misleading result. Furthermore, when investigators have compared individual and neighborhood-level SES, their dimensions can sometimes carry distinct associations with cognitive health (Cao et al., 2026; George et al., 2020). For example, in a seven-year follow-up study, individual and neighborhood dimensions of socioeconomic status played different, age-dependent roles in MCI risk (Fernández-Blázquez et al., 2021).

Research on place-based social determinants of health and late-life cognition has primarily focused on cognitive performance/decline, dementia risk, and biomarkers of AD/ADRD (L. M. Besser et al., 2017; Hunt et al., 2020; Krishnamurthy et al., 2025). However, very few studies have examined whether place-based SDOH measures are associated with impairment on cognitive screening measures, and the studies that have done so have been conducted in clinical or research settings (Liu et al., 2026; Rote et al., 2017). To our knowledge, no studies have examined how neighborhood SES and environmental burden are associated with performance on a cognitive screening measure that can be administered in community-facing settings. This gap is important because understanding what neighborhood living conditions or environmental features place people at greatest risk for dementia can help inform where health systems should direct dementia risk reduction efforts (Bergeron et al., 2022; Díaz-Santos et al., 2023). If the neighborhoods or other specific geographic areas where dementia risk is highest can be reliably identified, then it can inform where cognitive screening and other clinical services should be directed, expanding reach to underserved older adults who may face barriers to traditional diagnostic pathways (Tramujas Vasconcellos Neumann et al., 2026).

The present study examined the extent to which neighborhood SES and environmental burden were associated with cognitive impairment in a sample of community-dwelling older adults examined in community settings and compared these domain-specific associations against those of composite indices. We hypothesized that greater composite neighborhood disadvantage would be associated with greater odds of cognitive impairment. We also hypothesized that lower neighborhood SES and greater environmental burden would each be associated with higher odds of cognitive impairment.

## METHODS

### Participants

Eligible participants were community-dwelling adults between 50 and 89 years of age who resided in the state of Ohio. Individuals were excluded if they (a) had a prior diagnosis of dementia, (b) had visual or auditory impairments that would impede valid performance on SAGE, (c) could not read or understand English sufficiently to complete the self-report questionnaires, or (d) were otherwise unable to provide informed consent independently. Participants received financial compensation in the form of gift cards for completing the screening and the questionnaires.

Participants were recruited through various community engagement initiatives in Central Ohio from January 2025 to December 2025. A community advisory board with representatives from four local organizations (Central Ohio Chapter of the Alzheimer’s Association, African American Alzheimer’s and Wellness Association, African American Male Wellness Agency, and the Urban Aging Residents Coalition) was consulted to provide feedback on the study procedures and also collaborated with the research team to engage potential participants. Community events included delivering interactive seminars about brain health and modifiable dementia risk factors in-person (12 events) or on a virtual platform (5 events); administering cognitive screenings alongside other health screenings offered at local health fairs (6 events); and having a table at other events hosted by churches and other community organizations serving older adults (n = 7).

### Procedures

Participants had the option to either (a) complete all study components in person at community engagement events, (b) complete all study components online via a secure REDCap portal, or (c) complete the cognitive screening in person and complete the self-report questionnaires online at a later date. During in-person data collection, participants underwent informed consent with a trained research assistant. They then completed the SAGE and a series of self-report questionnaires on paper. After completion of the cognitive screening, SAGE assessments were scored and participants were provided individualized feedback regarding their results by trained study personnel. When screening results suggested signs of cognitive impairment, the study PI provided participants with information about getting a comprehensive neuropsychological assessment and other healthcare services for memory concerns. Participants completing the study online provided informed consent electronically and completed self-report questionnaires on REDCap. Next, they printed a copy of the SAGE from the REDCap portal, filled it out independently, and either (a) sent it back to the research team via postal mail, or (b) took a picture of each page and sent it via e-mail. A trained research assistant scored the SAGE and contacted the participant by phone to provide the results; the study PI directly contacted participants who scored below the cut-off for impairment to provide recommendations for pursuing additional healthcare services for memory concerns. All study procedures complied with the Declaration of Helsinki and were classified as exempt from full IRB review by The Ohio State University Behavioral and Social Sciences Institutional Review Board (STUDY20250685).

### Measures

#### Cognitive Function

Global cognitive function was assessed using the Self-Administered Gerocognitive Exam (SAGE), a self-administered cognitive screening measure designed to be sensitive to mild cognitive impairment. Participants were provided the test on four sheets of paper and read the instructions themselves, completing tasks that assess orientation, language, executive functioning, reasoning, and visuospatial abilities. Memory was assessed by a task on the last page that requires the participants to encode and recall a specific instruction from an earlier page. Total scores range from 0 to 22, with lower scores indicating poorer cognitive performance. In previous research, a cut-off of 17 or greater has been used to classify unimpaired individuals, 15-16 to denote individuals with mild impairment, and 14 or less to identify individuals with severe impairment.

#### Neighborhood metrics

Participants provided their home address or ZIP code + 4, as well as the length of residence in that particular neighborhood, which was used to generate neighborhood metrics. Four measures of neighborhood context were derived from the Environmental Justice Index (EJI), a national place-based index developed by the Centers for Disease Control and Prevention/Agency for Toxic Substances and Disease Registry (CDC, 2024). First was the EJI Overall Rank: a composite measure that integrates all of the socioeconomic, environmental, and health indicators within a particular census tract into a single index score. Second was the EJI Social-Environmental Rank (SER), which includes the census tract-level socioeconomic and environmental indicators but excludes the health vulnerability score, making it appropriate for research examining health outcomes without incorporating pre-existing disease burden into the exposure measure. Third was the EJI Social Vulnerability Rank, which isolates socioeconomic factors such as education attainment, poverty rates, home ownership, housing conditions, and transportation access at the census tract level. Fourth was the EJI Environmental Burden Rank, which isolates exposure to environmental hazards, including air pollution, proximity to toxic waste sites and transportation infrastructure that could contribute to pollution, as well as lack of access to health-promoting aspects of the built environment (e.g., recreational parks, walkability). EJI metrics are scored as ranks ranging from 0.00 to 1.00, where higher values denote greater social vulnerability (i.e., lower neighborhood-level SES), higher environmental burden, or worse local health conditions. In accordance with CDC/ATSDR guidelines, EJI percentile ranks were categorized into four qualitative groups using quartile cutoffs: Low (0.00–0.25), Low-Moderate (0.25–0.50), Moderate-High (0.50–0.75), and High (0.75–1.00). Supplemental Figure 1 displays the components that comprise the EJI overall rank and their hierarchical structure.

#### Statistical Analyses

All analyses were conducted using IBM SPSS Statistics 31. Descriptive statistics were used to characterize the sample, including means and standard deviations for continuous variables, and frequencies and percentages for categorical variables. Due to the low sample size of the mild impairment group (12.4%) relative to the other two groups (65.1% unimpaired, 22.4% severe impairment) in the present study, the mild and severe impairment groups were combined to form a single impaired group for all analyses (34.9%). SAGE impairment status was coded as 0 = unimpaired (SAGE ≥17) and 1 = impaired (SAGE ≤16). Income was recorded into four categories: $0–$29,999, $30,000–$74,999, $75,000 and over, and unknown income, with $0–$29,999 serving as the reference group in regression models. Independent-samples t-tests were used to compare impaired and unimpaired participants on continuous variables. Spearman correlations were used to assess associations among demographic characteristics, neighborhood metrics, and cognitive performance.

Hierarchical logistic regression analyses were conducted to examine the extent to which the neighborhood metrics measures predicted cognitive impairment status above and beyond demographic characteristics. Age, years of education, and income dummy variables were entered in Block 1, and each neighborhood-level predictor was entered separately in Block 2.

Four moderation analyses were conducted using PROCESS Model 1 to examine whether years of education moderated the associations between EJI measures and SAGE impairment status, controlling for age. Separate models were conducted for EJI Overall Rank, EJI-SER Rank, EJI Social Vulnerability Rank, and EJI Environmental Burden Rank. Statistical significance was evaluated using two-tailed tests with an alpha level of .05.

## RESULTS

### Demographic Characteristics

Table 1 displays the demographic characteristics and neighborhood metrics for the sample. The 281 participants were predominantly non-Hispanic Black (78.6%) and female (84.7%), with approximately one-third of whom performed in the impaired range on the SAGE. In comparison to the unimpaired participants, those classified as impaired were older (*t*(277) = -4.94, *p* < .001) and had fewer years of education on average (*t*(277) = 6.18, *p* < .001). Regarding neighborhood context, participants reported living in their current neighborhood for an average of 19.7 years (range = 2 months to 70 years). In the overall sample, all four EJI metrics were near the 50^th^ percentile based on national comparisons. EJI Overall and SER ranks were similar between unimpaired and impaired participants. Consistent with expectations, impaired participants also tended to have higher EJI Social Vulnerability Ranks (*M* = 0.5993, *SD* = 0.2847) than unimpaired participants (*M* = 0.4828, *SD* = 0.2785). In contrast, impaired participants had lower EJI Environmental Burden Rank scores (*M* = 0.4133, *SD* = 0.2456) than unimpaired participants (*M* = 0.4718, *SD* = 0.2452).

**Table 1.** Demographic characteristics at baseline for the overall sample (n = 281) and by impairment status.

| Variable | All Participants | Unimpaired (SAGE = 17 or more) | Impaired (SAGE = 16 or less) | Cohen's d |
| --- | --- | --- | --- | --- |
| **n (%)** | 281 (100.0%) | 183 (65.1%) | 98 (34.9%) |  |
| **SAGE Total Score** (mean ± SD (range)) | 17.4 ± 3.6 (6 – 22) | 19.6 ± 1.7 (17 – 22) | 13.3 ± 2.3 (6 – 16) | 3.22*** |
| **Age** (mean ± SD) | 67.7 ± 9.6 | 65.8 ± 9.2 | 71.4 ± 9.4 | -0.61*** |
| **Gender** (% women) | 84.7 % | 82.0 % | 89.8 % |  |
| **Race/Ethnicity** (%) |  |  |  |  |
| American Indian/Alaska Native | 0.4 % | 0.5 % | 0.0 % |  |
| Asian (non-Hispanic) | 1.4 % | 1.6 % | 1.0 % |  |
| Black (non-Hispanic) | 78.6 % | 77.6 % | 80.6 % |  |
| White (non-Hispanic) | 8.5 % | 12.6 % | 1.0 % |  |
| Mixed Race | 11.0 % | 7.7 % | 17.3 % |  |
| **Years of education** (mean ± SD) | 15.3 ± 2.3 | 15.9 ± 2.1 | 14.2 ± 2.3 | 0.76*** |
| **Educational attainment** (%) |  |  |  |  |
| Less than high school | 0.7 % | 0.5 % | 1.0 % |  |
| High school diploma/GED | 14.6 % | 5.5 % | 31.6 % |  |
| Some college/associate's degree | 30.2 % | 26.8 % | 36.7 % |  |
| Bachelor's degree | 26.3 % | 33.3 % | 13.3 % |  |
| Master's degree | 24.2 % | 29.0 % | 15.3 % |  |
| Professional/Doctoral degree | 3.9 % | 4.9 % | 2.0 % |  |
| **Employment (%)** |  |  |  |  |
| Employed | 28.5 % | 36.1 % | 14.3 % |  |
| Retired | 59.1 % | 55.2 % | 66.3 % |  |
| On Disability | 2.5 % | 2.7 % | 2.0 % |  |
| Unemployed | 0.7% | 0.5 % | 1.0 % |  |
| Unknown employment status | 1.4 % | 1.6 % | 1.0 % |  |
| **Income level (%)** |  |  |  |  |
| \$0 - \$29,999 | 16.7 % | 14.8 % | 20.4 % | |
| \$30,000 - \$74,999 | 33.5 % | 33.9 % | 32.7 % | |
| \$75,000 and over | 30.2 % | 36.1 % | 19.4 % | |
| Unknown income | 19.6 % | 15.3 % | 27.6 % |  |
| **Neighborhood Metrics (mean ± SD)** |  |  |  |  |
| EJI Overall Rank | 0.54 ± 0.32 | 0.51 ± 0.32 | 0.59 ± 0.31 | -0.22<sup>1</sup> |
| EJI Social-Environmental Rank | 0.52 ± 0.27 | 0.51 ± 0.27 | 0.54 ± 0.28 | -0.12 |
| EJI Social Vulnerability Rank | 0.52 ± 0.29 | 0.48 ± 0.28 | 0.60 ± 0.28 | -0.41** |
| EJI Environmental Burden Rank | 0.46 ± 0.25 | 0.48 ± 0.24 | 0.41 ± 0.25 | 0.26* |
| **Participation method (%)** |  |  |  |  |
| Completed study entirely in-person | 87.2% | 83.6% | 93.9% |  |
| Completed study entirely online | 7.5% | 11.5% | 0.0% |  |
| SAGE in-person, questionnaires online | 5.3% | 4.9% | 6.1% |  |
| **Community event type (%)** |  |  |  |  |
| In-person presentation | 37.4% | 38.3% | 35.7% |  |
| Health fair | 40.6% | 38.3% | 44.9% |  |
| Other tabling event | 11.7% | 8.2% | 18.4% |  |
| Virtual presentation / visited website | 10.3% | 15.3% | 1.0% |  |

**Note.** SAGE = Self-Administered Gerocognitive Exam.
\* $p < .05$
\*\* $p < .01$
\*\*\* $p < .001$
<sup>†</sup>Two-tailed $p = .069$

**Table 2:** Hierarchical Logistic Regression Model predicting SAGE Impairment Status by EJI Overall Rank.

| Predictors | B | p | OR | 95% Confidence Interval |  |
| --- | --- | --- | --- | --- | --- |
|  |  |  |  | Lower Bound | Upper Bound |
| Step 1 |  |  |  |  |  |
| Age | .058 | <.001 | 1.060 | 1.028 | 1.092 |
| Years of Education | -.313 | <.001 | .731 | .641 | .833 |
| Income (\$0 to \$29,999 vs. \$30,000 to \$74,999) | -.200 | .622 | .819 | .371 | 1.810 |
| Income (\$0 to \$29,999 vs. \$75,000 or more) | -.147 | .743 | .963 | .359 | 2.077 |
| Income (\$0 to \$29,999 vs. Unknown income) | .613 | .172 | 1.847 | .766 | 4.452 |
| Step 2 |  |  |  |  |  |
| EJI Overall Rank | .491 | .290 | 1.633 | .658 | 2.596 |

**Note:** B = unstandardized log-odds coefficient; 95% CI = 95% confidence interval. EJI = Environmental Justice Index.

**Table 3:** Hierarchical Logistic Regression Model predicting SAGE Impairment Status by EJI Social-Environmental Rank.

| Predictors | B | p | OR | 95% Confidence Interval |  |
| --- | --- | --- | --- | --- | --- |
|  |  |  |  | Lower Bound | Upper Bound |
| Step 1 |  |  |  |  |  |
| Age | .058 | <.001 | 1.060 | 1.028 | 1.092 |
| Years of Education | -.313 | <.001 | .731 | .641 | .833 |
| Income (\$0 to \$29,999 vs. \$30,000 to \$74,999) | -.200 | .622 | .819 | .371 | 1.810 |
| Income (\$0 to \$29,999 vs. \$75,000 or more) | -.147 | .743 | .963 | .359 | 2.077 |
| Income (\$0 to \$29,999 vs. Unknown income) | .613 | .172 | 1.847 | .766 | 4.452 |
| Step 2 |  |  |  |  |  |
| EJI Social-Environmental Rank | .094 | .861 | .910 | .319 | 2.596 |

**Note:** B = unstandardized log-odds coefficient; 95% CI = 95% confidence interval. EJI = Environmental Justice Index.

**Table 4:** Hierarchical Logistic Regression Model Including EJI Social Vulnerability Rank.

| Predictors | B | p | OR | 95% Confidence Interval |  |
| --- | --- | --- | --- | --- | --- |
|  |  |  |  | Lower Bound | Upper Bound |
| Step 1 |  |  |  |  |  |
| Age | .058 | <.001 | 1.060 | 1.028 | 1.092 |
| Years of Education | -.313 | <.001 | .731 | .641 | .833 |
| Income (\$0 to \$29,999 vs. \$30,000 to \$74,999) | -.200 | .622 | .819 | .371 | 1.810 |
| Income (\$0 to \$29,999 vs. \$75,000 or more) | -.147 | .743 | .963 | .359 | 2.077 |
| Income (\$0 to \$29,999 vs. Unknown income) | .613 | .172 | 1.847 | .766 | 4.452 |
| Step 2 |  |  |  |  |  |
| EJI Social Vulnerability Rank | 1.316 | .014 | 3.727 | 1.305 | 10.648 |

**Note:** B = unstandardized log-odds coefficient; 95% CI = 95% confidence interval. EJI = Environmental Justice Index.

**Table 5:** Hierarchical Logistic Regression Model Including EJI Environmental Burden Rank.

| Predictors | B | p | OR | 95% Confidence Interval |  |
| --- | --- | --- | --- | --- | --- |
|  |  |  |  | Lower Bound | Upper Bound |
| Step 1 |  |  |  |  |  |
| Age | .058 | <.001 | 1.060 | 1.028 | 1.092 |
| Years of Education | -.313 | <.001 | .731 | .641 | .833 |
| Income (\$0 to \$29,999 vs. \$30,000 to \$74,999) | -.200 | .622 | .819 | .371 | 1.810 |
| Income (\$0 to \$29,999 vs. \$75,000 or more) | -.147 | .743 | .963 | .359 | 2.077 |
| Income (\$0 to \$29,999 vs. Unknown income) | .613 | .172 | 1.847 | .766 | 4.452 |
| Step 2 |  |  |  |  |  |
| EJI Environmental Burden Rank | -1.302 | .025 | .272 | .087 | .846 |

**Note:** B = unstandardized log-odds coefficient; 95% CI = 95% confidence interval. EJI = Environmental Justice Index.

Figure 1 displays the distribution of participants by their EJI Overall Rank, EJI Social Vulnerability Rank, and Environmental Burden Rank; the SER ranks are not displayed because no accompanying map was available. EJI Overall and Social Vulnerability Ranks were distributed approximately evenly across the four qualitative categories. In contrast, the EJI environmental burden ranks were more heavily concentrated among individuals living in areas of moderate-high burden, followed by low burden.

**Figure 1.**
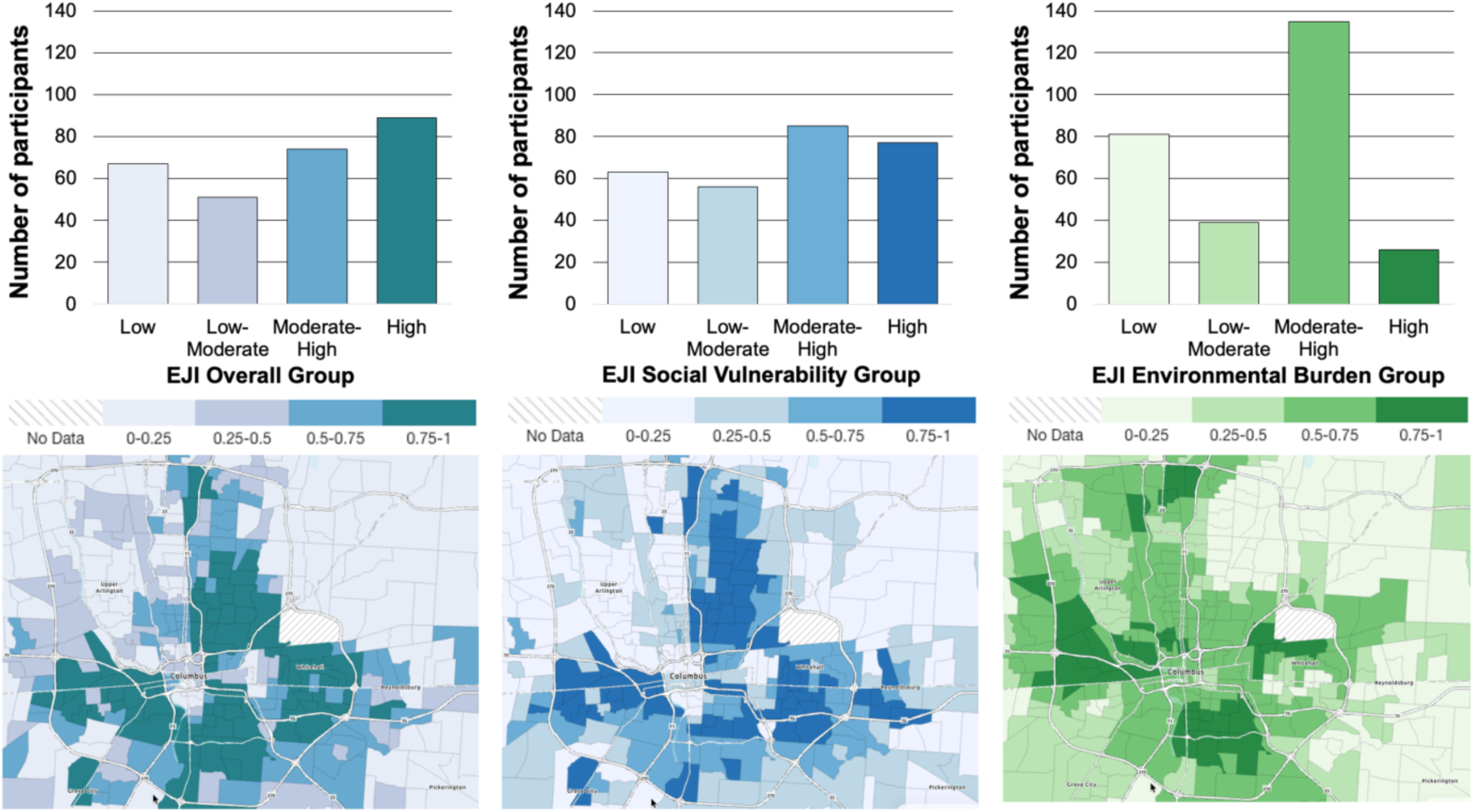
Geospatial distribution of the Environmental Justice Index (EJI) environmental burden and social vulnerability metrics across Ohio and Franklin County. Note. Maps display census tract-level EJI data for the State of Ohio with corresponding insets detailing Franklin County. Shading corresponds to index percentiles, where darker gradients indicate higher cumulative environmental burdens (green) and greater social vulnerability (blue). Percentile intervals range from 0.0-0.25 (lowest) to 0.75-1.0 (highest); diagonal-striped (hatched) regions indicate areas where data were unavailable due to a lack of residential population.

Figure 2 shows a cross-tabulation of census tract rankings that revealed significant structural divergence between neighborhood socioeconomic status and physical environmental burden. While traditional spatial epidemiology assumes that social deprivation and environmental pollution co-occur uniformly, these measures diverged in our sample. Notably, within the lowest tier of social vulnerability (high neighborhood-level socioeconomic status), 34.9% of participants nevertheless resided in tracts characterized by Moderate-to-High Environmental Burden, indicating that a substantial portion of the participants lived in urban neighborhoods with high economic resources and notable exposures to adverse environmental conditions.

**Figure 2.**
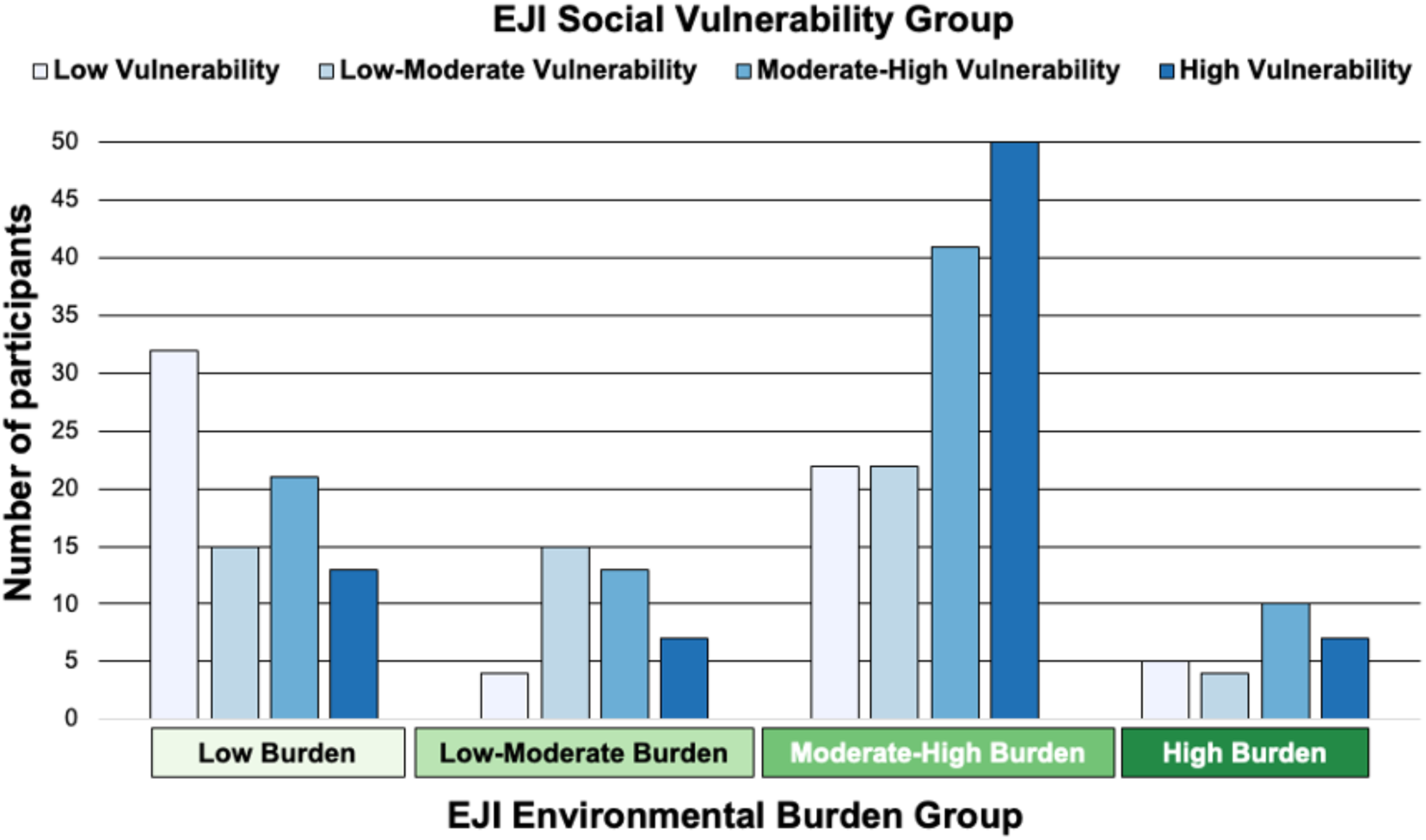
Cross-tabulation of participant counts by EJI Social Vulnerability and Environmental Burden categories.

**Figure 3.**
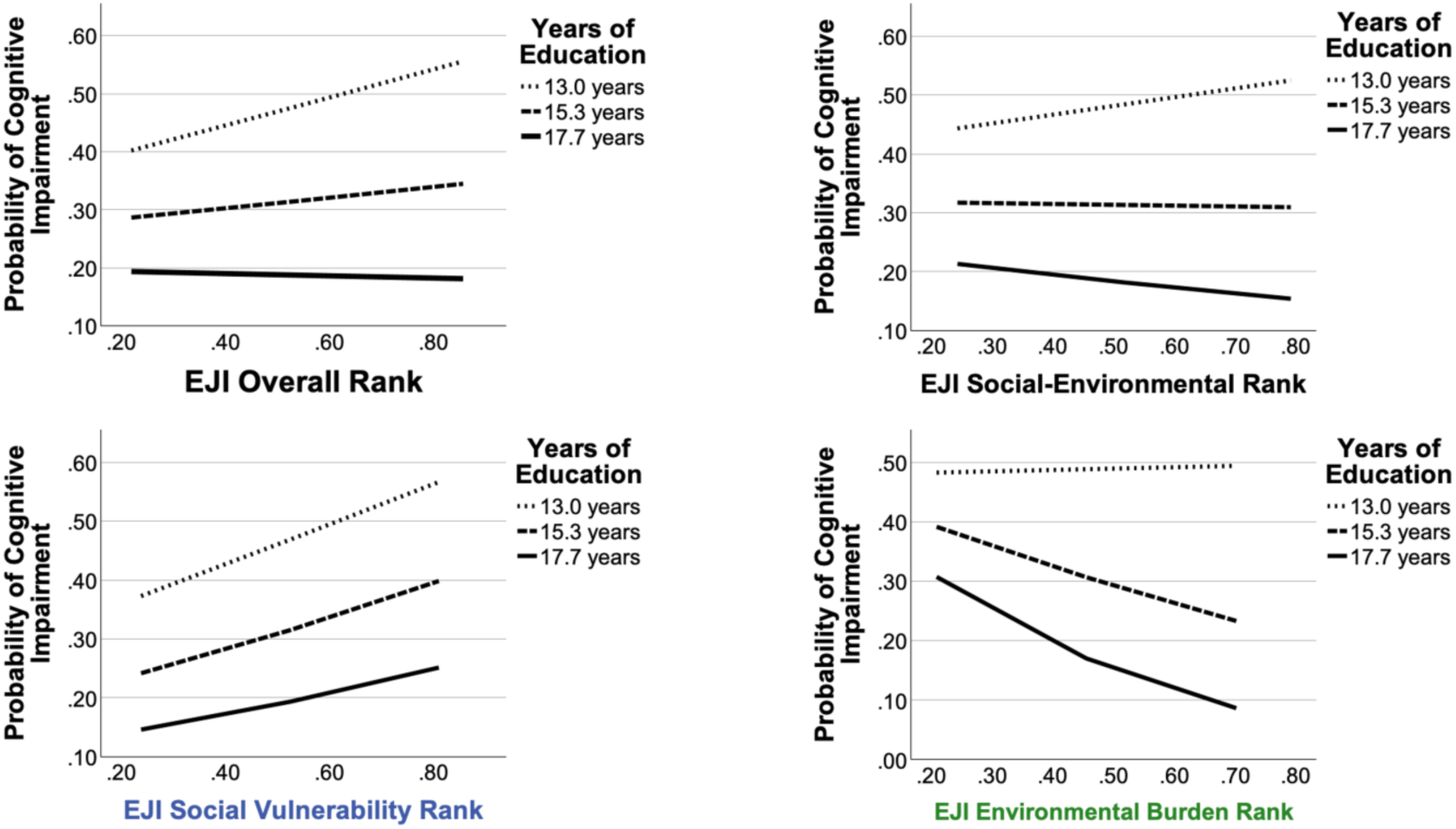
Simple slopes plots illustrating the moderation of Environmental Justice Index (EJI) effects on the likelihood of cognitive impairment by educational attainment. Note. Plots depict the moderated relationships between EJI metrics—Social-Environmental Rank (top panel), Social Vulnerability Rank (middle panel), and Environmental Burden Rank (bottom panel)—and the predicted log-odds of cognitive impairment (SAGE score ≤16). Simple slopes represent the conditional effects of each index across three distinct moderation levels of education, corresponding to the sample - 1.0 SD (13.0 years), mean (15.3 years; orange), and +1.0 SD (17.65 years; green). Estimates are derived from conditional logistic moderation models controlling for age.

### Hierarchical logistic regression predicting cognitive impairment status

As shown in Figure 2, the EJI Social Vulnerability Ranks appeared relatively evenly distributed across the four qualitative groups (low, low-moderate, moderate-high, and high vulnerability), whereas the Environmental Burden Ranks did not appear evenly distributed. To assess whether EJI Environmental Burden Ranks could be validly used as a continuous variable in a logistic regression, the linearity in the log-odds was formally assessed using the Box-Tidwell transformation procedure. The interaction between the continuous environmental burden rank and its natural log was non-significant (*B* = -1.41, *p* = .486), confirming that treating the EJI Environmental Burden Ranks as a continuous variable was statistically valid.

In the covariate block, age, years of education, and income variables significantly predicted cognitive impairment status, χ²(5) = 55.30, *p* < .001, with a Nagelkerke *R*² of .246. The covariate model correctly classified 73.3% of cases. Older age was associated with greater odds of impairment, OR = 1.06, 95% CI [1.03, 1.09], *p* < .001, whereas more years of education was associated with lower odds of impairment, OR = 0.73, 95% CI [0.64, 0.83], *p* < .001.

In the next block of the hierarchical regression, the addition of the EJI Overall rank did not significantly improve model fit, χ² change(1) = 1.125, p = .289. The full model remained statistically significant, χ² (6) = 56.438, p < .001, with a Nagelkerke R² of .251 and overall classification accuracy of 75.8%. In the full model, EJI-SER Rank was not significantly associated with impairment status after controlling for age, years of education, and income level, B = -.491, OR = 1.63, 95 % CI [0.77,4.45], p = .172.

Similarly, in a separate hierarchical regression, the addition of the EJI-SER rank after the covariates did not significantly improve model fit, χ² change(1) = 0.031, p = .861. The full model remained statistically significant, χ² (6) = 55.33, p < .001, with a Nagelkerke *R*² of .246 and overall classification accuracy of 73.3%. EJI-SER Rank was not significantly associated with impairment status in the full model, *B* = -0.094, OR = 0.91, 95 % CI [0.32, 2.60], *p* = .861.

In contrast to the EJI Overall Rank and the EJI-SER, the addition of EJI Social Vulnerability Rank did significantly improve model fit, χ² change (1) = 6.18, *p* = .013. The full model was statistically significant, χ²(6) = 61.48, *p* < .001, with a Nagelkerke *R*² of .271 and overall classification accuracy of 75.4%. In the full model, greater EJI Social Vulnerability scores were significantly associated with higher odds of impairment, *B* = 1.316, OR = 3.73, 95% CI [1.31, 10.65], *p* = .014.

The addition of EJI Environmental Burden Rank also significantly improved model fit, χ² change(1) = 5.17, *p* = .023. The full model was statistically significant, χ²(6) = 60.47, *p* < .001, with a Nagelkerke *R*² of .267 and overall classification accuracy of 72.6%. However, in contrast to the EJI Social Vulnerability Rank, EJI Environmental Burden Rank was inversely associated with impairment, *B* = -1.302, OR = 0.27, 95% CI [0.09, 0.85], *p* = .025.

### Moderation analyses

Four moderation analyses using PROCESS Model 1 were conducted to determine whether the relationship between each of the four neighborhood metrics and SAGE impairment status was moderated by years of education. Age was included as a covariate and was significantly associated with SAGE impairment status in all four models (*p* < .001).

For EJI Overall Rank, the full model was statistically significant, *p* < .001, Nagelkerke R² = .264. In line with the results from the hierarchical regression, EJI Overall Rank was not significantly associated with SAGE impairment status in the moderation model, *B* = 4.04, *p* = .181. The interaction between EJI Overall Rank and years of education was not statistically significant, *χ*²(1) = 1.38, *p* = .241; therefore, years of education did not significantly moderate the relationship between EJI Overall Rank and SAGE impairment status.

For EJI-SER Rank, the full model was statistically significant, *p* < .001, Nagelkerke *R*² = .231. EJI-SER Rank was not significantly associated with SAGE impairment status in the moderation model, *B* = 4.28, *p* = .236. Years of education did not significantly moderate the relationship between EJI-SER Rank and SAGE impairment status, *χ*²(1) = 1.39, *p* = .238.

For EJI Social Vulnerability Rank, the full model was statistically significant, *p* < .001, Nagelkerke *R*² = .252. EJI Social Vulnerability Rank was not significantly associated with SAGE impairment status in the moderation model, *B* = 1.93, *p* = .559, and years of education did not significantly moderate the relationship between EJI Social Vulnerability Rank and SAGE impairment status, χ²(1) = 0.04, p = .848.

In contrast to the other three models, EJI Environmental Burden Rank was associated with SAGE impairment status *B* = 9.09, *p* = .030. Furthermore, the interaction between EJI Environmental Burden Rank and years of education was statistically significant, χ²(1) = 6.16, p = .013, indicating that years of education moderated the relationship between EJI Environmental Burden Rank and SAGE impairment status.

Conditional effects were examined to interpret the significant EJI Environmental Burden Rank × years of education interaction. At 13.0 years of education, EJI Environmental Burden Rank was not significantly associated with SAGE impairment status. At 15.3 years of education, EJI Environmental Burden Rank was significantly associated with lower odds of impairment, *B* = -1.52, *p* = .010. At 17.7 years of education, EJI Environmental Burden Rank was significantly associated with lower odds of impairment, *B* = -3.13, *p* = .002. These findings indicate that the inverse association between EJI Environmental Burden Rank and SAGE impairment status was most evident among participants with higher years of education.

## DISCUSSION

This study examined the extent to which neighborhood socioeconomic status and environmental burden were associated with impaired performance on a cognitive screening measure in a sample of urban older adults. Overall, the findings add nuance to a wealth of research showing that neighborhood-level social determinants of health may play a key role in shaping late-life cognitive health; under certain circumstances, different features of a particular neighborhood may have diverging or even opposing associations with cognitive health.

Consistent with our hypothesis, poorer neighborhood-level socioeconomic status was associated with higher odds of impairment on the SAGE. This finding remained true after controlling for age, education, and income level. Neighborhood socioeconomic status may influence cognitive aging through multiple pathways. Beyond the effects of individual-level income and financial stability, older adults who lived in neighborhoods with low economic resources may be exposed to cumulative stressors such as high housing cost burdens, limited digital connectivity, and lower access to high-quality healthcare. Economic strain and housing instability can induce chronic stress responses that compromise vascular and neural health over time (Low et al., 2025; Sol et al., 2022). Simultaneously, healthcare access barriers can hinder the proactive management of cardiometabolic disease (Islam et al., 2022). Together, these social conditions can compound over time to elevate the risk of cognitive decline and dementia.

Contrary to our expectations, higher environmental burden was associated with lower risk of cognitive impairment; individuals living in neighborhoods with worse environmental hazards tended to perform better on the SAGE than those in neighborhoods with fewer environmental hazards. Crucially, this observed inverse association should be interpreted with caution and not as evidence that environmental hazards are neuroprotective. A key explanation for this unexpected finding lies in the spatial overlap between neighborhood-level socioeconomic status and exposure elements of the built environment that may be hazardous. As demonstrated by the cross-tabulation of the neighborhood metrics, only a small portion of the participants lived in areas of high environmental burden, and few lived in areas of low-to-moderate environmental burden. The majority lived in areas of low or moderate-to-high environmental burden, approximating a bimodal distribution. Furthermore, over half of the participants residing in low environmental burden census tracts also lived in areas of moderate-to-high social vulnerability, whereas nearly a third of those in moderate-to-high burden tracts belonged to low-vulnerability tiers. Consequently, without conditioning on individual characteristics such as education or income level, the unmoderated baseline model confounds lower physical hazard exposure with underlying structural social deprivation. The apparent neuroprotective effect of environmental burden may therefore be an artifact of urban demographic tendencies rather than a true buffer against cognitive decline.

Another potential explanation is that performance on the SAGE may be more heavily influenced by factors associated with or sensitive to neighborhood-level socioeconomic status (e.g., poverty, lack of internet access) than features of the built environment. In addition, the significant moderating role of education likely reflects unmeasured behavioral factors—such as daily activity patterns—where higher-educated adults can more readily spend daytime hours in less burdened environments despite living in higher-burden tracts. Notably, the expected inverse relationship between environmental burden and cognitive performance did emerge among participants with lower levels of education (∼13 years). Overall, the counterintuitive finding regarding environmental burden and cognitive impairment underscores the necessity of decomposing multi-dimensional neighborhood metrics and accounting for individual reserve mechanisms when evaluating cognitive health. Notably, the composite measures used to evaluate the total effect of neighborhood context—the EJI Overall Rank and EJI-SER—did not significantly predict impairment status in adjusted models. Given that the measure of neighborhood socioeconomic status and the measure of hazards in the built environment had diverging associations with cognitive impairment, the composite measures may have masked the distinct relationships observed when the domains were examined separately in this sample. These findings suggest that domain-specific neighborhood measures may provide more meaningful information than combined indices when examining cognitive impairment risk.

Exploratory analyses revealed that individual years of education altered the relationship between neighborhood environmental burden and cognitive impairment. Crucially, the counterintuitive inverse association described earlier—where residing in areas with worse environmental burden was associated with lower odds of cognitive impairment—was driven almost exclusively by highly educated older adults. Albeit an unexpected finding, this pattern aligns directly with the framework of cognitive reserve (Stern et al., 2020). Higher educational attainment may provide a buffer against cognitive decline, enabling individuals to maintain strong performance on cognitive screening measures even when the cumulative effects of neighborhood environments would predict poorer cognitive performance. In this sample, highly educated older adults may frequently reside in dense, commercial, or transit-heavy areas that are in close proximity to aspects of the built environment that could have adverse health effects. For example, participants in our sample who lived in suburban areas near the international airport may have higher levels of education and classify as living in a high neighborhood-level SES census tract. However, these same individuals have higher exposure to air pollution due to their proximity to the airport and thereby a worse EJI environmental burden. Because these individuals possess significant cognitive reserve, their intact performance on the cognitive screening may contribute to the observed inverse association, possibly creating the finding that neighborhood physical hazards are benign or even protective. However, for individuals with lower formal education, this apparent protection is no longer evident. Higher educational attainment may simply mask the overt impacts of exposure to environmental hazards on a cognitive screening tool, rather than neutralizing the underlying biological risks associated with air pollution or high-traffic transportation infrastructure (Weuve et al., 2021). The findings align with a growing body of research showing the importance of integrating macro-level neighborhood metrics with individual characteristics to better elucidate how neighborhood context impacts late-life cognitive health (Allan et al., 2022) .

This study has several limitations. First, the cross-sectional design prevents conclusions about causality or directionality. Although neighborhood-level measures were associated with cognitive impairment status, it cannot be determined whether neighborhood conditions contributed to cognitive impairment over time. Second, cognitive impairment was assessed using the SAGE, a screening tool rather than a clinical diagnosis. While SAGE is useful for community-based screening and has been validated for detecting mild cognitive impairment and early dementia, future studies should incorporate clinical diagnoses, comprehensive neuropsychological assessments, and/or longitudinal follow-up to confirm cognitive outcomes (Scharre et al., 2021). Furthermore, the sample was almost entirely urban and relatively well-educated (15.3 years of education on average); only one participant lived in a high commuting area according to Rural-Urban Commuting Area (RUCA) codes, and only two participants completed less than a high school diploma. Therefore, the findings regarding the moderating effect of education may not be as applicable to older adults living in rural areas, individuals with low levels of education, or other underrepresented groups.

The unexpected inverse association between environmental burden and impairment status may reflect unmeasured factors or limitations in how the ‘neighborhood’ was defined in this study. Using a person’s home address and census tract as a proxy of their exposure to neighborhood hazards may not capture the environments where participants spend most of their daytime hours. For example, individuals may live in higher-burden areas but work, receive services, socialize, or spend substantial time in other neighborhoods. In addition, because we captured their home address at a single timepoint, we cannot fully account for the cumulative effects of neighborhood-level exposures to factors that may promote or hinder brain health across the lifespan. Furthermore, given that this sample was predominantly Black (78.6%; all of African American heritage, except for one Afro-Latino and one East African participant), another important dimension of neighborhood context is residential segregation. A wealth of research has demonstrated that living in an area of high current or historical racial residential segregation is associated with greater dementia risk among Black older adults, but not necessarily among older adults from other racial/ethnic groups (Fisk et al., 2026; Meyer et al., 2023). Previous studies indicate that composite measures of neighborhood context and other social determinants of health can yield diverging associations with health outcomes that diverge between racial groups, despite lacking direct indicators of segregation; this occurs because such composite measures inherently incorporate the upstream and downstream consequences of residential segregation (M. S. Wong et al., 2023).

Despite these limitations, the present findings highlight the importance of considering neighborhood context in cognitive aging research. The association between social vulnerability and impairment status suggests that cognitive screening and dementia prevention efforts may benefit from attention to broader structural and social conditions, beyond individual factors. Additionally, community-based cognitive screening tools such as SAGE may be especially useful for reaching older adults who face barriers to traditional diagnostic pathways (Scharre et al., 2014). Accessible community-based screening may help reduce barriers to early dementia detection in underserved populations (Jin et al., 2025). These findings also suggest that domain-specific social determinants of health measures may be more informative than combined indices when examining cognitive impairment risk. Given that the EJI Environmental Burden Rank captures multiple environmental indicators, future research could examine more precise associations between these individual indicators and cognitive health to identify specific components that are most relevant to dementia risk. Future research should use longitudinal designs, more precise environmental exposure measures, and larger samples to clarify how neighborhood socioeconomic and environmental conditions interact with individual-level resources to shape cognitive aging.

Overall, these findings suggest that the neighborhood context may offer valuable information for predicting risk for cognitive impairment among older adults. Importantly, neighborhood socioeconomic status and built environment may have different effects on cognition, and these effects are influenced by individual-level characteristics that may provide a buffer against cognitive decline. These results highlight and support the need to examine both individual and contextual determinants of cognitive health to better understand and address disparities in cognitive impairment among older adults.

## Data Availability

All data produced in the present study are available upon reasonable request to the authors

## ACKNOWLEDGEMENTS

This work was supported by a grant from the Alzheimer’s Association (AARFD-24-1309508). The authors used a secure enterprise version of the Gemini generative AI model (Google) accessed via The Ohio State University to assist with editing the manuscript and generation of SPSS syntax. All study design, data analysis decisions, verification of statistical results, interpretation of findings, and final manuscript content were reviewed and approved by the authors.

## CONSENT STATEMENT

All human participants provided informed consent prior to participation in this study.

